# Does TB Vaccination Reduce COVID-19 Infection? No Evidence from a Regression Discontinuity Analysis

**DOI:** 10.1101/2020.04.13.20064287

**Authors:** Masao Fukui, Kohei Kawaguchi, Hiroaki Matsuura

## Abstract

In the middle of the global COVID-19 pandemic, the BCG hypothesis, the prevalence and severity of the COVID-19 outbreak seems to be correlated with whether a country has a universal coverage of Bacillus-Calmette-Guérin (BCG), a vaccine for tuberculosis disease (TB), has emerged and attracted the attention of scientific community and media outlets. However, all existing claims are based on cross-country correlations that do not exclude the possibility of spurious correlation. We merged country-age-level case statistics with the start/termination years of BCG vaccination policy and conducted a regression discontinuity and difference-indifference analysis. The results do not support the BCG hypothesis.

## I. Introduction

There is no vaccine against the rapidly spreading novel Corona virus disease (COVID-19), which has contributed to more than 1.2 million infections and 70,000 deaths worldwide. In search of the effective prevention and treatment for COVID- 19, some researchers found that there was a strong correlation between the heath effect of COVID-19 and national policy for Bacille Calmette-Guerin (BCG), a vaccine for tuberculosis (TB) disease. Miller et al. (2020) found that both morbidity and mortality due to COVID-19 are associated with early adoption or universal coverage of BCG vaccination, while Giovanni Sala and Tsuyoshi Miyakawa (2020) found that BCG vaccination slowed down the spread or progression of symptoms rather than reduced COVID-19 death. Shet et al. (2020) also examine the correlation with mortality. In the middle of global pandemic, the BCG hypothesis has quickly attracted the attention of popular media outlets such as Bloomberg (March-30-2020) and New York Times (April-5-2020). Another study by Zhang et al. (2013) further indicates that the BCG strain with fewer epitopes deleted, such as the Japan and Russian/Bulgarian strain, are more likely to be effective due to higher and more frequent responses, although this paper is not about COVID-19. Clinical trial for these BCG strains are now initiated in several countries (For example, see Medical News Today (April-2-2020)).

These previous findings are based on the cross-country association between health outcomes of COVID-19 and the national BCG vaccination policies, thus do not exclude the possibility of spurious correlation. For example, a country with higher BCG vaccination coverage is more likely to be poor as infectious diseases (such as TB) are still leading causes of death. Such country is less likely to be connected to the major economic regions such as China, Europe, and the United States due to their trade openness and geographic location. There is an imminent need to re-examine this newly emerging hypothesis for two reasons. If there is no possibility, the society should spend more effort searching for more plausible solutions to combat COVID-19. Moreover, if people demand BCG vaccination for COVID-19, this creates a shortage for BCG vaccination for children who actually need it. In fact, Japan BCG Laboratory, the only producer of BCG vaccine in Japan, shipped three times more vaccine in March 2020 than the average in the past.^1^ The Japan Society for Vaccinology recently warned that BCG vaccination was for preventing TB, not for other purposes in their official statement.^2^

We believe that clinical trials can provide us with a secure basis for the effectiveness of BCG vaccination against COVID-19, but it is worth reexamining this purported hypothesis using a more credible identification strategy before spending more time and resources to test it. In this paper, we test this hypothesis with a best available identification strategy based on observational data: a regression discontinuity and difference-in-difference analysis.

## II. Materials and Method

We collected information on year of introduction and termination of the BCG vaccination and types of strains used from the BCG World Atlas (Zwerling et al., 2011). Then, we matched them with country-age-level COVID-19 confirmed case statistics from information available from each government’s website. This resulted in a data set over 17 countries with relevant information: Australia, Colombia, the Czech Republic, Denmark, Finland, India, Japan, Korea, Latvia, New Zealand, Romania, Singapore, Spain, Sweden, Switzerland, Thailand, and Vietnam. Exact age-level case statistics were available in Colombia, the Czech Republic, Thailand, Vietnam, and Singapore. In remaining countries, only a 10-year age-group level data were available. Of these 17 countries, the Japan strain or Russia/Bulgaria strain have been used in Japan, Thailand, Colombia, and Latvia. In Colombia, case statistics at the nationality level are available. Hence, we focus on people with Colombian nationality in Colombia data, whereas all residents not differentiated by nationality are included in the other countries’ data. The immunization rate of infants at each year is available from the World Health Organization (WHO) website.^3^ We refer to this in the figure, even though this does not necessarily correspond to the immunization rate at each age as of 2020.

We first conducted a regression discontinuity analysis using data from Colombia, the Czech Republic, Thailand, Vietnam, and Singapore. The identification assumption is that factors such as basic hygiene that can affect the COVID-19 infection rate do not discontinuously change around the age at which the BCG vaccination was introduced. If a comprehensive health care reform is implemented simultaneously with the introduction of a universal BCG policy, then this assumption is violated. However, any bias should occur in the same direction with the expected BCG effects, producing an upward bias. In this paper, we demonstrate that even with this potential upward bias, we do not observe any improvement in the COVID-19 infection rate at the age of policy change.

In Colombia, the BCG vaccination was introduced in 1960 as a mass campaign targeted at young people under 15 years old (Arbeláez, Nelson and Muñoz, 2000). In 1978, the strain changed from English/Japan to French. Thus, we can study the effects at three distinct time boundaries: 1945 (i.e. 1960 minus 15) when people covered by the campaign were born, 1960 when the vaccination was introduced, and 1978 when the strain changed. In the Czech republic, BCG vaccination was introduced in 1953 for people under 18 (Tu et al., 2012), and stopped universal vaccination in 2010 (Vaśáková, 2013). They changed from the Plague to the Russia strain in 1981 and changed from the Russia to the Danish strain in 1994. This paper investigated the effects at 1935 (i.e. 1953 minus 18), 1953, 1981, 1994, and 2010. In Thailand, the BCG vaccination was introduced in 1977 for infants. In 1987, they changed from the Danish to the Japan strain. Between 1987 and 1991, they revaccinated at age 7. We study the effects at 1977 and 1991. In Vietnam, the BCG vaccination was introduced in 1985 for infants (Jit et al., 2015). Older people were not vaccinated, because they do not have a revaccination policy. In Singapore, the BCG vaccination started in 1957 (Goh, 1985). Because they revaccinate at ages 6, 11, and 15, we also studied the effect at 1942 (i.e. 1957 minus 15). Vietnam and Singapore do not use the Japan or Russia/Bulgaria strains.

Second, we conducted a difference-in-difference analysis using all 17 countries with 10-year-age-group level case statistics. The identification assumption in the current model is that without the BCG vaccination the expected log difference of the infection rates across age-groups are the same across countries. In this paper, we constructed a treatment variable that indicates the age ratio within age-group *t* covered by the BCC vaccination (*BCG*_*it*_) in country *i*. We also constructed a treatment variable that indicates the vaccination using the Japan or Russia/Bulgaria strain (*BCG*_*JRB,it*_) and another treatment variable that indicates the vaccination using other strains (*BCG*_*Other,it*_). We then regressed the log of the number of cases per thousand on these treatment variables, controlling for country and age-group dummies as:

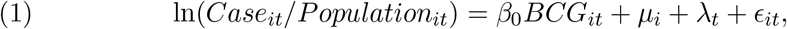

or:

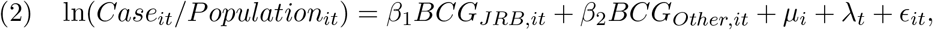

where *µ*_*i*_ and *λ*_*t*_ are country *i* and age-group *t* specific fixed effects. We ignored the effects of potential coverage by revaccination policy in the difference-in-difference analysis.

## III. Results

Figure 1 displays the results of the regression discontinuity analysis for Colombia, the Czech Republic, Thailand, Vietnam, and Singapore. The figures from Vietnam and Singapore indicate that the immunization rate quickly increased after the vaccination policy. Thus, the discontinuity of treatment at the policy changes seems to be justified. In (a) Colombia, there is no significant decrease after coverage (74 years old at the end of 2019), the start of vaccination policy (59 years old): nor is there a significant increase after the change of strains (41 years old). In (b) the Czech Republic, there is no significant decrease after coverage (84 years old), the start of vaccination policy (66 years old), the change to the Japan strain (38 years old): nor is there a significant increase after the change from the Japan strain (25 years old), and the termination of the vaccination policy (10 years old). In (c) Thailand, there is no decrease after the start of vaccination policy (42 years old) or after the change from Danish to Japan strain (32 years old). In (d) Vietnam, there is no significant drop after coverage and start of vaccination policy (34 years old). In (e) Singapore, there is no significant decrease after coverage (77 years old), and the start of the vaccination policy (62 years old). In summary, results from regression discontinuity analysis do not support either the positive BCG effects, or the stronger BCG effects with the Japan and Russia/Bulgaria strains. Some argue that the effect of BCG lasts only for 20 to 30 years and hence would not be detected at old cohorts. However, the apparent no effect at the age of termination of the universal vaccination policy in the Czech Republic in 2010 would address this issue.

**Figure 1.**
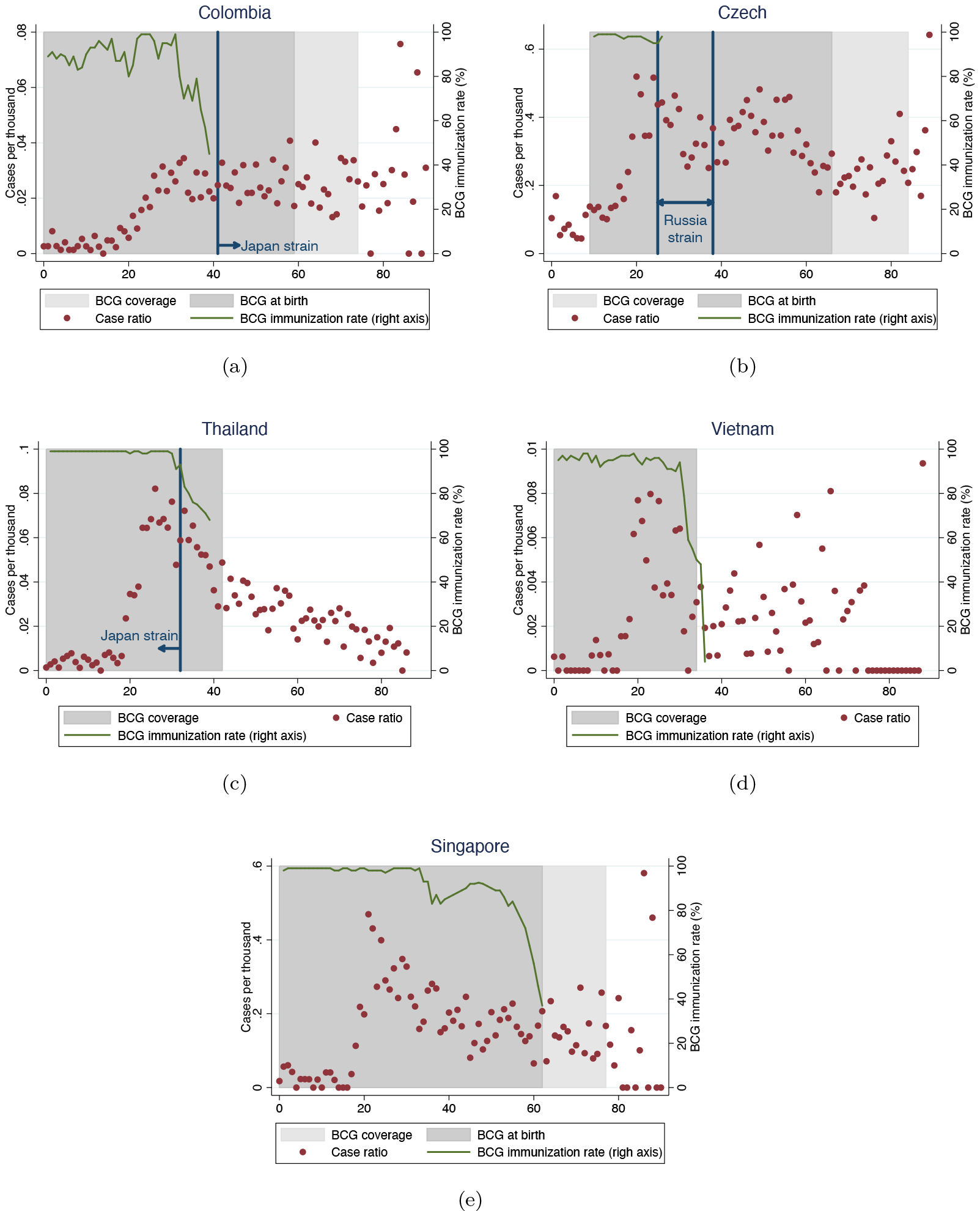
Regression Discontinuity Analysis. Note: BCG coverage is the age at which a vaccination policy targeted. BCG at birth is the period when a universal BCG vaccination policy exists. The immunization rate is plotted only when available.

Figure 2 displays the results of the difference-in-difference analysis. The Figure’s left panel shows the scatter plot of residualized log cases per thousand and the residualized BCG coverage ratio after controlling for country and age-group dummies from Model (1). If the BCG reduces COVID-19 infection, we should expect a negative slope: however, here the slope is positive, indicating that our results do not support the purported BCG effects on COVID-19 infection. The right panel of Figure 2 shows the scatter plot of residualized log cases per thousand and the residualized BCG coverage with the Japan or the Russia/Bulgaria strains. The highlighted circles represent Japan, Thailand, Colombia, and Latvia where the Japan or Russia/Bulgaria strains were once used. Again, we do not observe a negative slope: therefore, our results do not support the hypothesis that the Japan and the Russia/Bulgaria strains that are close to the original Tuberculosis are especially effective. This difference-in-difference analysis includes cohorts for which the universal BCG vaccination coverage terminated during 1990s and 2000s. Thus, the result also addresses the problem that potential BCG effects last only for 20 to 30 years. Note that when we remove the country fixed effects, the sign of the coefficient becomes significantly negative, consistent with the previous cross-country analysis of Miller et al. (2020). This underscores the importance of controlling for unobserved country-specific characteristics.

**Figure 2.**
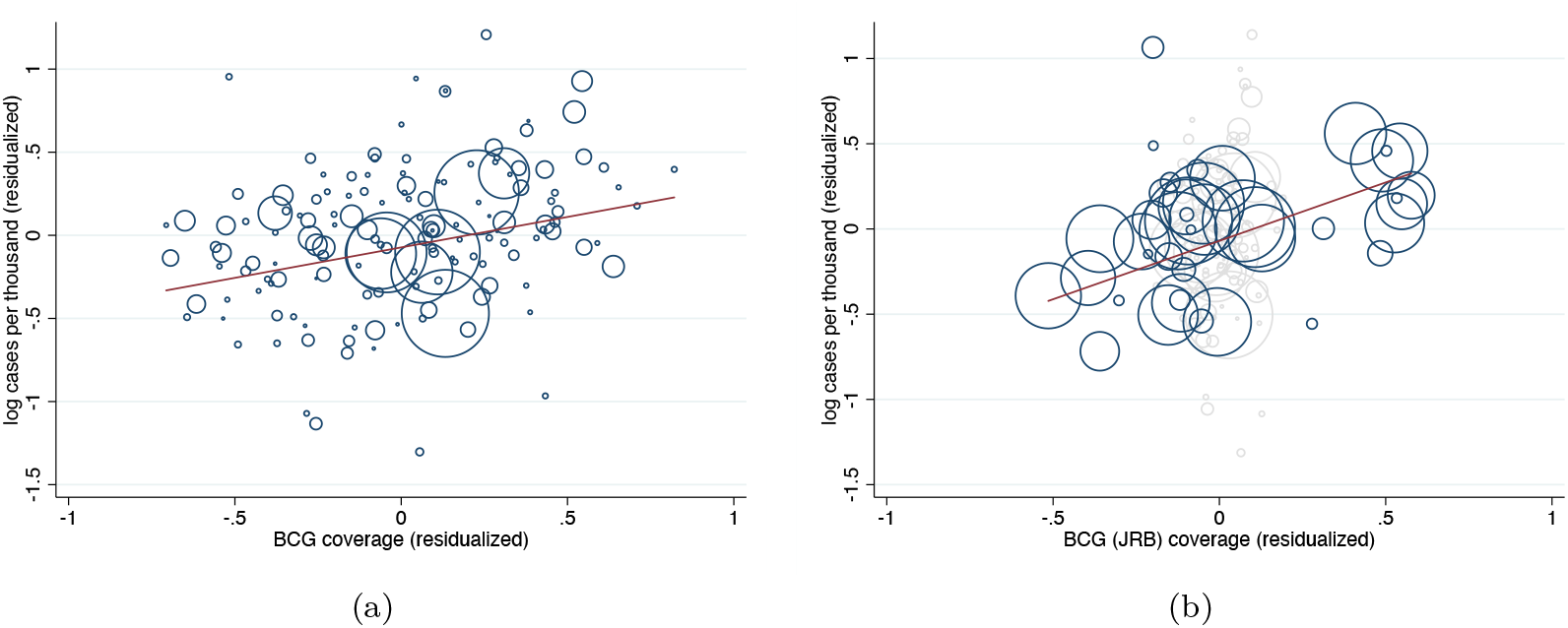
Difference-in-Difference Analysis. Note: The sample is at the country and 10-years age-group level. The y-axis is the log cases per thousands after controlling for country and age dummies. The x-asis is the ratio of ages covered by BCG vaccination within each age group after controlling for country and age dummies. The left panel is based on Model (1) and the right panel is based on Model (2). In the right panel, countries with Japan or Russia/Bulgaria strains are highlighted. The size of each circle represents the population size, and the solid line corresponds to a regression line weighted by the population size.

## IV. Discussion

There are three limitations to our analysis. First, our analysis focused on the effect on COVID-19 infection due to the data availability. BCG immunization can still prevent from developing symptoms or reduce death once infected by COVID-19. However, given that those with COVID-19 symptoms and their close contacts are more likely to be tested and confirmed, we still capture some effects of BCG immunization on developing symptoms. Second, there is possibility that population (in addition to individual) immunity affects our COVID-19 outcomes. However, we removed out potential population immunity effects of the BCG vaccination by controlling for country-specific unobserved fixed effects. We agree that population immunity effects are another parameter of policy interest. However, we leave this to the future research. Another potential criticism is that people acquired immunity from the actual infection of TB, rather than the BCG immunization. If the effects of the BCG immunization and TB on the COVID-19 infection are the same and the TB infection rate was almost 100% around the change in the vaccination policy, there is a possibility that we failed to pick up any additional effects of the BCG immunization. However, this is implausible. Marks et al. (2018) document that in Vietnam, only around 30-40% of age 30s and 40-50% of older population are infected by TB in 2016. In Thailand, the infection rate of children under 14 years old in 1977, when the vaccination started, was 15.2% (Sriyabhaya et al., 1993). Since Thailand and Vietnam are among the highest TB burden countries in the sample we study,^4^ we view these numbers as upper-bounds. There is also possibility that actual immunization rate did not reflect policy change, but this is also implausible given that BCG vaccination coverage rapidly increased from around 0 to 100% when a universal vaccination policy was introduced, as in Vietnam. Hence, if any BCG-specific effect exists, it should appear at the age of policy change.

## Data Availability

Data and replication codes are available at https://github.com/kohei-kawaguchi/BCG_replication

## Footnotes

1 Nish-Nihon Shinbun, accessed, April 8, 2020, https://www.nishinippon.co.jp/item/n/599010/ (in Japanese)

2 Japan Society for Vaccinology, accessed on April 3 2020, http://www.jsvac.jp/pdfs/kenkai.pdf

3 Reported estimates of BCG coverage, accessed on April 7 2020, https://www.who.int/data/gho/data/indicators/indicator-details/GHO/bcg-immunization-coverage-among-1-year-olds-(-). We add pre-1980 data for Singapore from Goh (1985).

4 WHO estimates, https://www.who.int/tb/country/data/download/en/.

